# SARS-CoV-2 Delta variant saliva viral load is 15-fold higher than wild-type strains

**DOI:** 10.1101/2021.11.29.21266980

**Authors:** Kenichi Imai, Ryo Ikeno, Hajime Tanaka, Norio Takada

**Author notes:** Correspondence: Kenichi Imai, D.D.S., Ph.D., Professor and Chairman, Department of Microbiology, Nihon University School of Dentistry, 1-8-13 Kanda-Surugadai, Chiyoda-ku, Tokyo 101-8310, Japan.

## Abstract

The emergence of SARS-CoV-2 Delta variants has escalated COVID-19 cases globally due to their high transmissibility. Since saliva is crucial for SARS-CoV-2 transmission, we hypothesized that a higher viral load of Delta variants in saliva than their parental wild-type strains contributed to the high transmissibility in the first place. However, studies have not reported this particular comparison done with viral copy numbers. Twenty-two genetically confirmed -positive saliva samples for wild-type strain and 32 Delta variants were statistically compared for viral copy number per milliliter determined by real-time qPCR combined with synthesized viral RNA and Poisson’s null distribution equation between the groups of wild and variant strains and between whole saliva and centrifugal supernatant in each group. We found that the copy number of the Delta variants was 15.1 times higher than wild-type strains of the whole saliva. In addition, the viral load of both strains in the whole saliva was higher than the pertinent supernatant, indicating that most viruses in the whole saliva are associated with host cells. Meanwhile, more than a million virions per milliliter of the viral load of the variants in the supernatants were 4.0 times higher but not significant than wild-type strains. Humanity must share our findings; the simple but concrete note that Delta variant viral load is abundant in the saliva is critical for preventing the spread of infection.

## INTRODUCTION

Since coronavirus disease 2019 (COVID-19) was officially declared a pandemic by the World Health Organization, severe acute respiratory syndrome coronavirus 2 (SARS-CoV-2) has killed over 5.1 million people (1). Emergence of SARS-CoV-2 Delta variants has escalated COVID-19 cases globally, which is probably due to its high transmissibility (2).

Saliva is crucial for SARS-CoV-2 transmission in three routes: droplets, contact, and aerosol infections. Moreover, saliva contains infectious virions released from oral epithelia and salivary glands (3). We have hence hypothesized that a higher viral load of Delta variants in saliva than their parental wild-type strains contributes to their high transmissibility in the first place. However unexpectedly, to the best of our knowledge, studies have yet reported this kind of comparison done with viral copy number.

## METHODS

To compare the viral load, we utilized whole saliva specimens collected from individuals visiting a fever outpatient clinic at Nagoya, Japan, at different time points before and after the emergence of Delta variant (November and December, 2020 and August and September, 2021, respectively). Detection of virus-positive saliva samples and L452R mutation was performed by one of major clinical laboratories, Bio Medical Laboratories Inc (BML) Tokyo, Japan. To determine viral load as viral copy number, extraction of viral RNA and real-time quantitative polymerase chain reaction (qPCR) were conducted referring to a method provided by the National Institute of Infectious Diseases of Japan (4). Simultaneously, using synthetized standard viral RNA, RNA levels were measured using Poisson’s null distribution equation. Comparisons of viral copy number between the variants and wild-type strains were determined statistically (Mann–Whitney’s *U*-test or Wilcoxon’s signed-rank test). Viral localization in saliva, i.e., whether virions are associated with host cells or floating away from them, was investigated by comparing the viral load between whole saliva and supernatant samples after centrifugation (10000 × g, 5 min). Additionally, the proportion of host cells isolated through the centrifugation was revealed with human chromosome-specific qPCR to be 99.5%, and it was not found in the supernatant (data not shown).

## RESULTS

Saliva samples from 22 wild-type strain-positive individuals (12 males, 39.5 ± 18.6 years of age and 2.6 ± 1.4 days since onset of symptoms) and 32 L452R mutation-positive individuals (20 males, 32.4 ± 14.8 years old and 3.6 ± 2.6 days since onset) were collected. There were no significant differences in average age (*p* = 0.12), gender (*p* = 0.57), or average days since onset (*p* = 0.096) between the two groups (Student’s *t*-test). Delta variants’ median copy number (1.86E+7, IQR: 3.16E+6∼8.03E+7) was 15.1 times significantly higher than wild-type strains (1.23E+6, 1.76E+5∼2.70E+7) per milliliter of whole saliva (Mann–Whitney’s *U*-test: *p* = 0.020) (Fig. 1). Although this is the first study determining Delta variant’s load in saliva, the viral copy number correlated well with C_T_ value determined by BML (Spearman’s rank correlation coefficient: *r*_*s*_ = −0.872, *p* = 8.71E-18) (Fig. 2). In addition, viral load of both variants and wild-type strains in whole saliva was significantly higher (15.9 times and 4.2 times, respectively; Wilcoxon’s signed-rank test: *p* = 1.2E-5 and *p* = 6.6E-3, respectively) than pertinent supernatant (Fig. 1), indicating that most viruses in whole saliva are associated with host cells. Meanwhile, the viral load of the variants in the supernatants was 4.0 times higher (1.17E+6, 1.40E+5∼3.31E+6) but not significant (Mann– Whitney’s *U*-test: *p* = 0.18) than wild-type strains (2.96E+5, 4.61E+4∼2.39E+6).

**Fig. 1.**
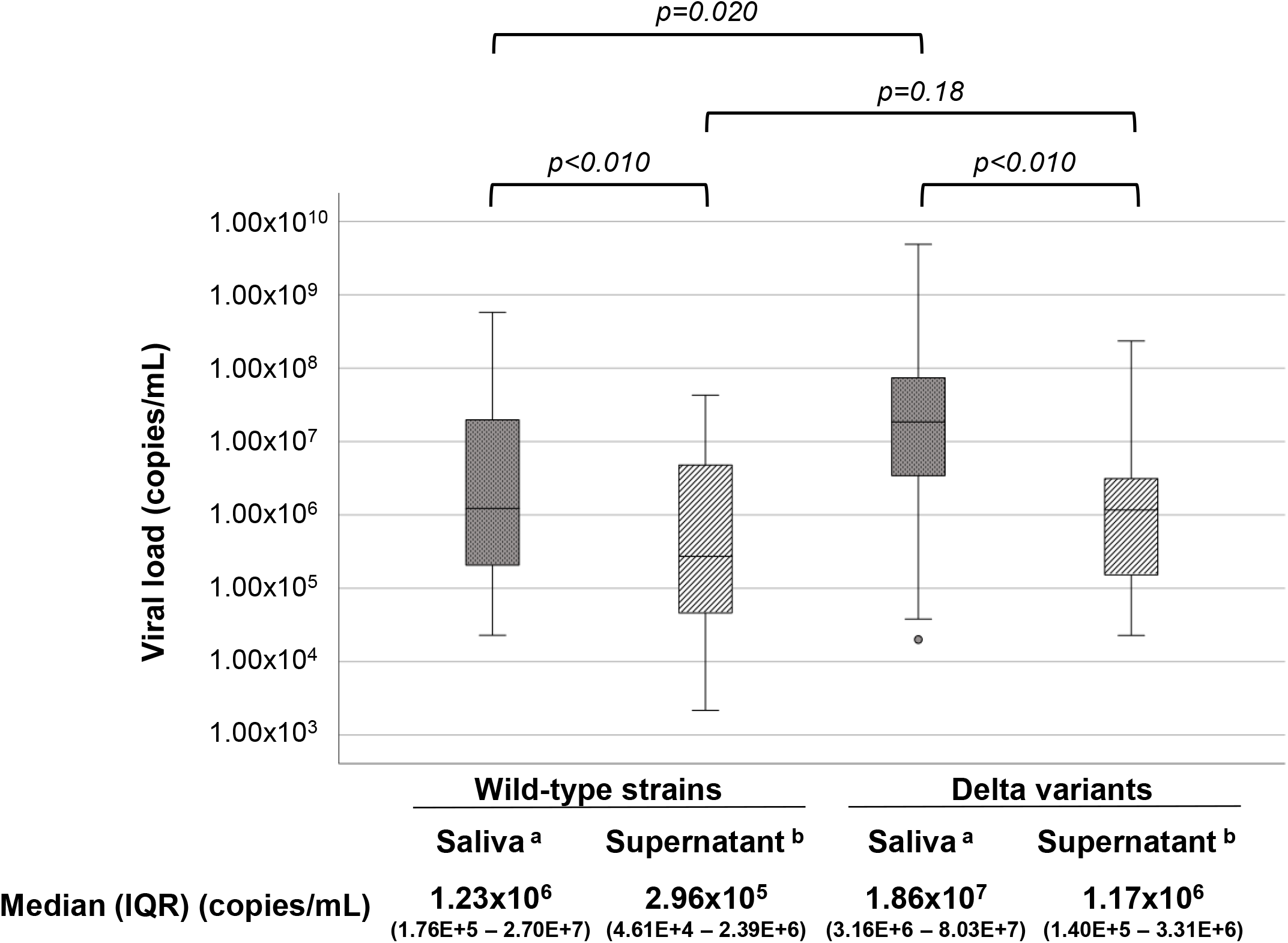
Comparisons between wild-type and Delta variant strains in whole saliva and its centrifugal supernatant. Comparisons between wild-type strains and Delta variants and between whole saliva and pertinent supernatant were determined using Mann–Whitney’s *U*-test and Wilcoxon’s signed-rank test, respectively. Viral load was significantly lower in saliva supernatant than whole saliva for both strains (*p* = 1.2E-5 for Delta variants and *p* = 6.6E-3 for wild-type strains). Delta variant’s viral load was higher than the wild-type strain in whole saliva (*p* = 0.020). Viral loads of both variants and wild-type strains in whole saliva were significantly higher (15.9 times and 4.2 times, respectively; *p* = 1.2E-5 and *p* = 6.6E-3, respectively) than the pertinent supernatant. Delta variant’s viral load in the supernatant was 4.0 times higher (1.17E+6) but not significant (*p* = 0.18) than wild-type strains (2.96E+5). Data are presented as median (IQR). a, whole saliva b, saliva centrifugal supernatant (10000 × g, 5 min)

**Fig. 2.**
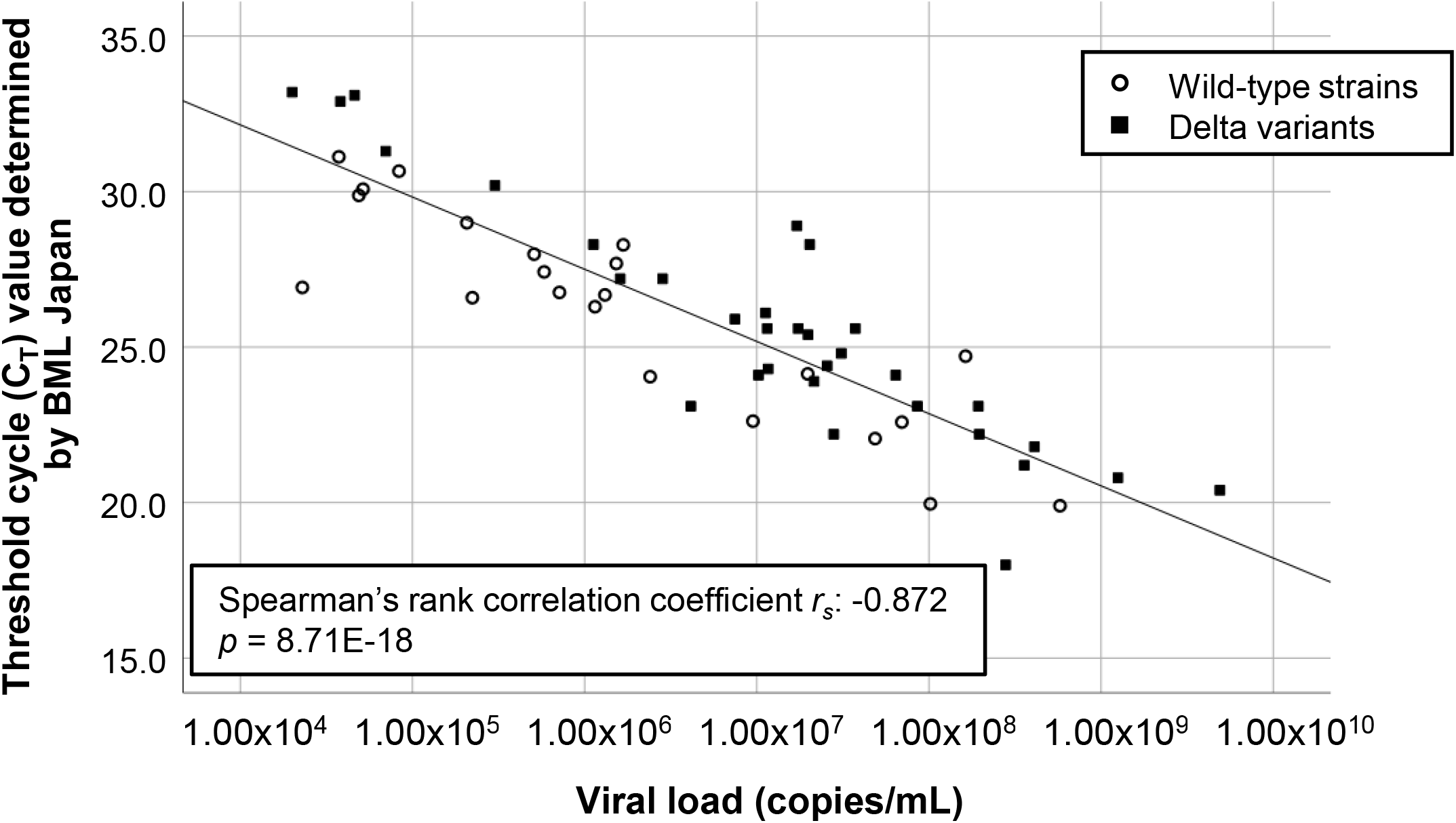
Spearman’s rank correlation analysis between the current study values (viral copies/mL) and BML laboratory results (C_T_ values) For both wild-type strains and Delta variants in whole saliva; the viral copy numbers we determined correlated with the C_T_ values analyzed by the BML laboratory.

## DISCUSSION

Our findings imply that the variants are excreted into saliva 15 times more than the wild-type strains, suggesting that droplets released from variant-infected individuals contain more virions. Moreover, as shown above, the variants’ median number in the supernatant 1.17E+6 virions per milliliter suggests that aerosols generated by variant-infected individuals contain more than one million virions per milliliter. This may also explain why Delta variant escalated COVID-19 cases globally because the basic reproduction number approximately 5.0 of the Delta variant is comparable with that of varicella virus, which is more contagious and transmitted not only by droplet but also airborne infections (5, 6). We believe that it is imperative for humanity to share our findings; the simple but concrete note that Delta variant viral load is abundant in saliva is critical for preventing the spread of infection.

## Data Availability

All data produced in the present study are available upon reasonable request to the authors

